# Modelling the impact of adopting new-generation insecticide-treated nets on malaria transmission and insecticide resistance

**DOI:** 10.64898/2026.03.05.26347588

**Authors:** Hamenyimana E. Gervas, Maranya M. Mayengo, Frank Chacky, Yeromin P. Mlacha, Halfan S. Ngowo, Fredros O. Okumu, Prashanth Selvaraj

## Abstract

**Background:** The widespread insecticide resistance increasingly threatens malaria elimination, prompting a reassessment of vector control strategies. As Tanzania transitions from standard pyrethroid-only insecticide-treated nets (ITNs) to new-generation nets, evaluating the impact of this shift on malaria transmission and resistance is critical.

**Methods:** Using the agent-based malaria model, EMOD, we assessed the impact of three ITN types, standard pyrethroid-only nets, pyrethroid-PBO nets (Olyset® Plus/PermaNet® 3.0), and the dual active, Interceptor® G2 nets (IG2) on malaria transmission and the evolution of insecticide resistance. We also evaluated different sequences for introducing the new-generation nets, and the impact of combining ITNs with indoor residual spraying (IRS). The model was calibrated using incidence and prevalence data from two regions in northwestern Tanzania, incorporating seasonality, insecticide resistance, and behaviors of dominant vectors *Anopheles funestus* (highly anthropophilic, endophilic) and *Anopheles arabiensis* (more opportunistic readily biting non-human hosts outdoors).

**Results:** Changing from standard pyrethroid-only ITNs to pyrethroid-PBO and thereafter to IG2 ITNs reduced homozygous-resistant *An. funestus* and *An. arabiensis* by 62.2% and 92.8%, respectively, and reduced incidence and prevalence by 94% and 75.2% respectively, under conditions where the probability of mosquito pyrethroid resistance was 0.75. Deploying IRS before the peak malaria transmission season in mid-May, in the second year following pyrethroid-PBO ITNs distribution, and repeating this every three years, reduced malaria incidence and prevalence by 76.4% and 52%, respectively.

**Conclusion:** In contrast to continuous use of standard pyrethroid-only ITNs, which sustains resistance selection, transitioning to new-generation ITNs, with or without periodic IRS, can disrupt the evolutionary trajectory of pyrethroid resistance, reduce malaria burden, and strengthen progress toward elimination.

## Background

Malaria vector control has expanded significantly since 2000, primarily through insecticide-treated nets (ITNs) and indoor residual spraying (IRS) as core insecticide-based interventions [1]. Since their inception, these interventions, together with improved case management, have prevented an estimated 2.2 billion malaria cases and 12.7 million deaths [2,3]. Despite being preventable and treatable, malaria remains a major global health challenge, with over 90% of the burden reported in sub-Saharan Africa [4,5], where the recent increase in malaria cases is driven in part by rising resistance to antimalarial drugs [6,7] and insecticides [8].

In particular, widespread insecticide resistance, reported in over 87% of malaria-endemic countries, poses a major obstacle to malaria elimination [4,9]. The dual use of pyrethroids in agriculture and public health has been a major driver of insecticide resistance [10–13]. In response, the World Health Organization (WHO) has endorsed next-generation insecticide-treated nets that combine pyrethroids with either an enzyme synergist or a second insecticide with an alternative mode of action [14]. Examples of these new generation ITNs include pyrethroid-piperonyl butoxide (PBO) nets (e.g. Olyset® Plus, containing permethrin and PBO, and PermaNet® 3.0, containing deltamethrin and PBO) as well as dual-active ingredient nets such as Interceptor® G2 (IG2 ITNs), which combines the pyrethroid alpha-cypermethrin with the pyrrole chlorfenapyr [9,14].

In Tanzania, insecticide resistance especially to pyrethroids has been detected in 22 of the 62 vector surveillance sentinel sites and confirmed in all major malaria vectors (*Anopheles gambiae sensu stricto (s*.*s*.*), Anopheles arabiensis*, and *Anopheles funestus*), with resistance in these species primarily mediated not only by metabolic detoxification enzymes but also multiple target site mutations [15–18]. Persistent resistance remains one of the major barriers to achieving the national target of reducing malaria incidence and mortality by at least 90% by 2030 [19]. In response to this challenge, the National Malaria Control Programme (NMCP) has previously implemented non-pyrethroid IRS using organophosphate or neonicotinoid insecticides in few councils in the lake zone, replacing the previously used pyrethroid IRS [19–21]. However, following the recent discontinuation of IRS [22,23], the NMCP, in collaboration with the US President Malaria Initiative (PMI) and Global Fund (GF), has been distributing pyrethroid-PBO ITNs (Olyset® Plus and PermaNet® 3.0) in areas previously reliant on IRS and standard pyrethroid-only ITNs mainly through a targeted mass campaigns every three years; while also deploying IG2 ITNs in few districts with the highest levels of resistance [22,23].

Despite ongoing initiatives, the impact of transitioning to pyrethroid-PBO and IG2 ITNs on malaria transmission and resistance dynamics, particularly in areas previously covered by standard pyrethroid-only ITNs and IRS, remains insufficiently understood. While some modelling studies have examined the impact of standard pyrethroid-only and next-generation ITNs [24–28], many overlook key factors such as seasonality, species-specific dynamics, resistance selection and the role of intervention rotation and transition. This stresses the urgent need for in-depth research to evaluate their impacts on both transmission and resistance evolution and propagation.

This study used an agent based mathematical model of malaria transmission, EMOD [24], to simulate the impact of transitioning from standard pyrethroid-only ITNs to new-generations ITNs (pyrethroid-PBO, and IG2) on malaria incidence and prevalence, and on the evolution and propagation of insecticide resistance. The study considers malaria transmission by *An. funestus* and *An. arabiensis*, the two dominant vectors in Tanzania, along with varying insecticide resistance profiles and seasonality.

## Methods

### EMOD malaria transmission model

In this study, all simulations were performed using the agent-based mechanistic model of malaria transmission (EMOD v2.20) [24,25], which represents individual humans and mosquitoes and their interactions. The model integrates all four developmental stages of mosquito species: eggs, larvae, pupae and adult mosquitoes, as local mosquito population dynamics are primarily driven by the availability of larval habitats [29]. The availability of larval habitats at any given time determines the number of larvae each habitat type can support, which consequently influences the number of adult vectors that emerge at the end of the life cycle [30,31]. Moreover, insecticide resistance is modeled as a phenotypic trait determined by allele combinations in mosquitoes. These combinations are represented through homozygous and heterozygous genotypes [25], to illustrate the selection dynamics of resistant mosquitoes and the propagation of resistance under ITN transition versus repeated deployment of the same insecticides. It is important to clarify that the primary focus of this study is on how different phenotypic resistance profiles influence mosquito selection through genetic changes and affect the effectiveness of ITNs. Detailed information on the mosquito life cycle, insecticide resistance mechanisms, malaria transmission modelling and genotype-phenotype interactions, including gene flow patterns in EMOD can be found in previous publications [24,25,32].

### Characterization of mosquito species

In Tanzania, *Anopheles gambiae* s.s., *An. arabiensis*, and *An. funestus* are primary malaria vectors. Although *An. gambiae* s.s. contributes to malaria transmission in some settings [33], widespread insecticide use, particularly through ITNs, has shifted vector composition, with *An. arabiensis* and *An. funestus* now dominating in many regions [33–37]. Given their current epidemiological significance and resistance profiles [19,35,38], this study modeled vector populations using *An. arabiensis* and *An. funestus*, assigning indoor feeding/resting fractions of 63.1% and 78.2% [39], respectively, with indoor feeding and resting assumed equal. These species were further assigned anthropophilic indices of 62% and 84%, respectively [39,40]. Seasonality was modeled based on observed monthly variations in mosquito abundance from lake zone (northwestern) in Tanzania [36].

### Model calibration

In Tanzania, ITNs serve as the primary malaria intervention and have been widely distributed nationwide since 2007 [41]. Since then, significant efforts have been made to enhance ITN coverage through various distribution channels, including mass campaigns, school net programs, antenatal care and targeted replacement campaigns [41]. Following the cessation of IRS activities in 2021, the country has been undergoing a phased transition, beginning with the replacement of standard pyrethroid-only ITNs with pyrethroid-PBO ITNs, followed by the deployment of IG2 ITNs in selected districts [23] (Refer to Figure 1 for the timeline of ITNs and IRS).

**Figure 1:**
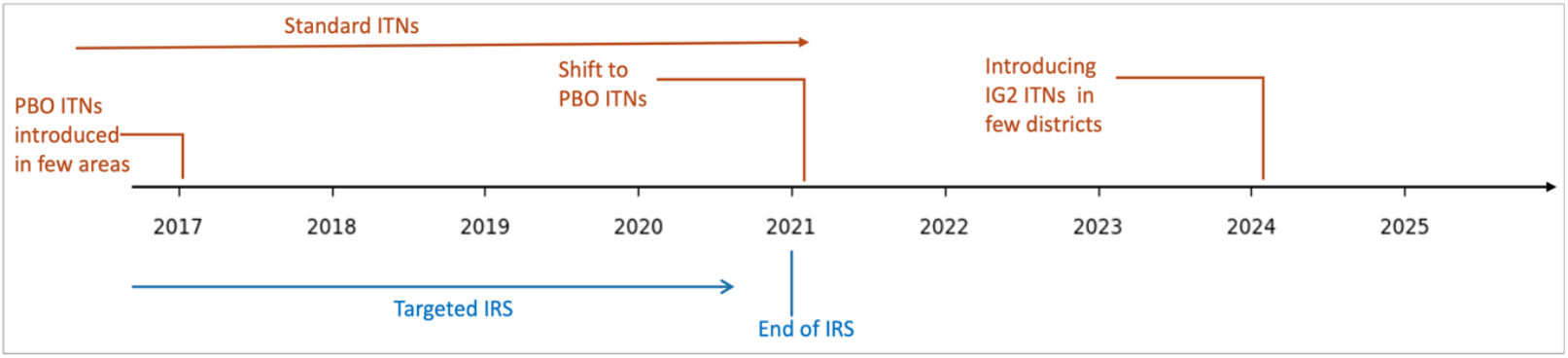
Timeline of ITN and IRS interventions in Tanzania (2017-2025) illustrating the transition from standard pyrethroid-only ITNs (standard ITNs) to new-generation ITNs [19,42].

The model was calibrated using the 2020 monthly prevalence data from five councils in Geita Region, Tanzania (Bukombe, Chato, Mbogwe, Nyang’hwale, and Geita District Councils) [43], and the 2019 monthly incidence data from four councils of Mwanza region (Misungwi, Ukerewe, Magu and Kwimba) [44]. These councils represent the lake zone of Tanzania, characterized by widespread insecticide resistance, prior IRS implementation, and the ongoing transition to new-generation nets. The datasets informed the adjustment of monthly larval habitat parameters for *An. funestus* and *An. arabiensis* to reproduce observed transmission patterns. For each larval habitat parameter set, simulations were compared with field data using mean squared error (MSE), and the best fit was obtained by averaging the curves with the lowest errors, resulting in a MSE of 0.0008 (Figure 2). During calibration, a cohort of 1,000 individuals was simulated over 40 years to establish age-specific immunity. The incidence dataset was not stratified by age, whereas the prevalence data specifically represented children aged 0-5 years. The model reproduces the observed seasonal dynamics, capturing the peak malaria transmission (Figure 2).

**Figure 2:**
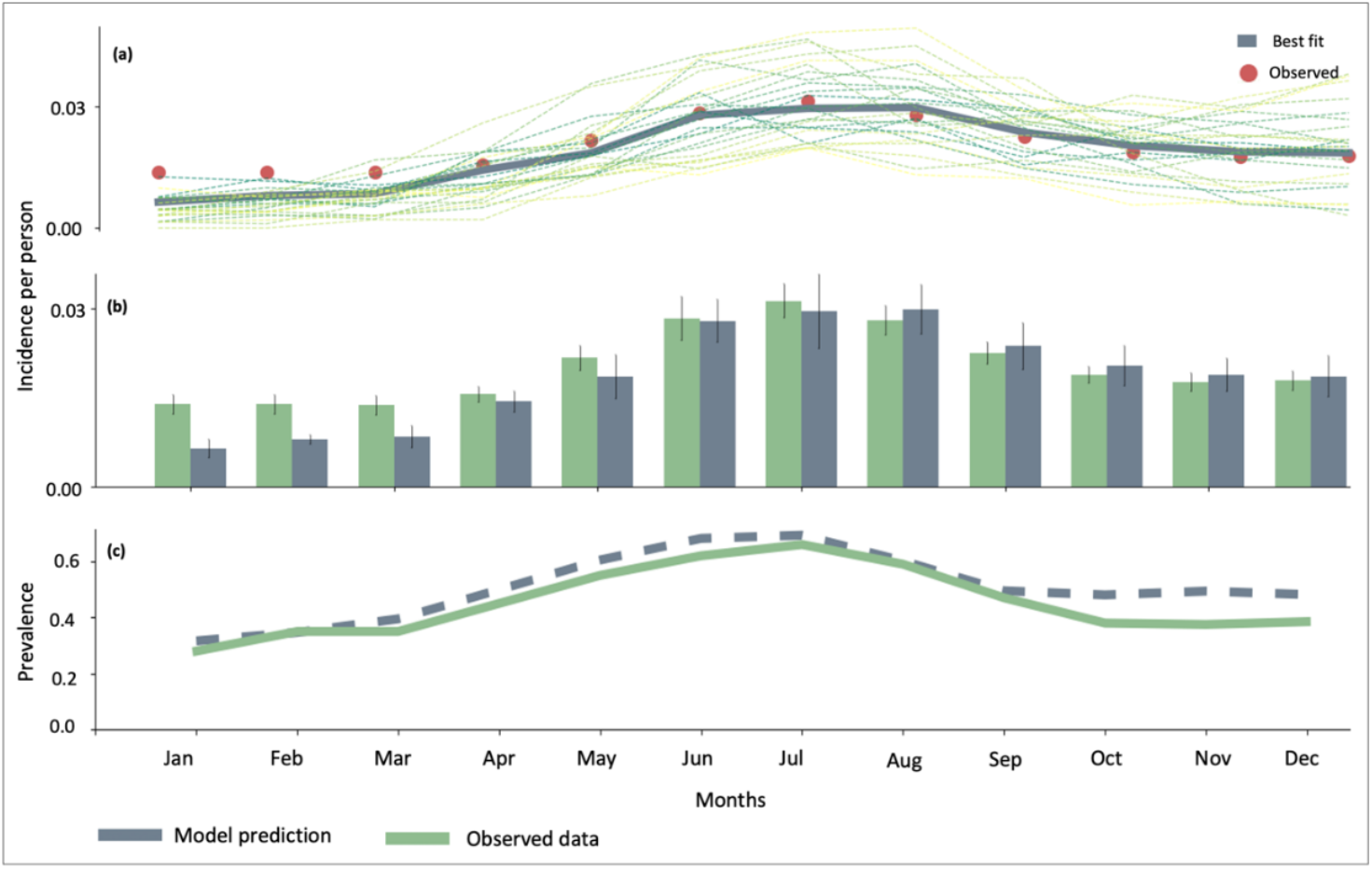
Model calibration: **(a)** the dashed lines represent model realizations with relatively low mean squared errors, the grey curve indicates the best-fit; **(b)** best fit versus observed incidence cases; **(c)** model prediction versus observed prevalence. The incidence data were calibrated using 2019 data from the four districts of Mwanza region [44], while prevalence was calibrated using 2020 data from five districts of Geita region [43].

### Modelled interventions

This study models the impact of standard-pyrethroid only, pyrethroid-PBO and IG2 ITNs and combinations of ITNs with organophosphate or neonicotinoid-based IRS. No additional interventions were considered. All the candidate interventions target adult female mosquitoes during host-seeking and resting, with uniform coverage set at 80%. ITNs are distributed on a triennial basis, typically preceding the onset of the peak malaria transmission season in March (Figure 2). However, this illustration may not accurately represent current practices in Tanzania, as the precise timing of ITN deployment is not well defined. See Table 1 in Appendix 1 for descriptions of all the parameters.

**Table 1:**
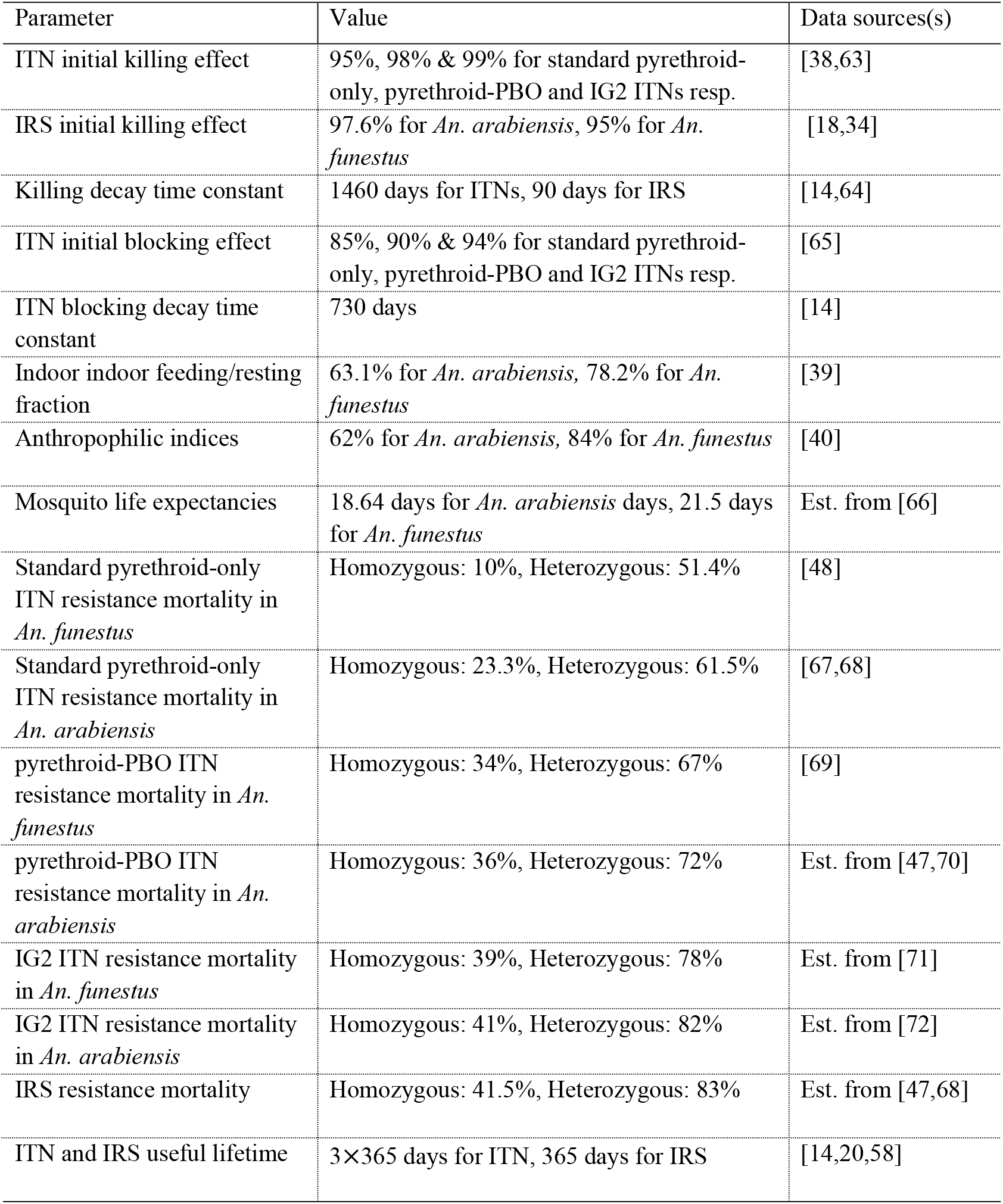
Key parameters and values used for simulations. Est=Estimated.

| Parameter | Value | Data sources(s) |
| --- | --- | --- |
| ITN initial killing effect | 95%, 98% & 99% for standard pyrethroid-only, pyrethroid-PBO and IG2 ITNs resp. | [38,63] |
| IRS initial killing effect | 97.6% for *An. arabiensis*, 95% for *An. funestus* | [18,34] |
| Killing decay time constant | 1460 days for ITNs, 90 days for IRS | [14,64] |
| ITN initial blocking effect | 85%, 90% & 94% for standard pyrethroid-only, pyrethroid-PBO and IG2 ITNs resp. | [65] |
| ITN blocking decay time constant | 730 days | [14] |
| Indoor indoor feeding/resting fraction | 63.1% for *An. arabiensis*, 78.2% for *An. funestus* | [39] |
| Anthropophilic indices | 62% for *An. arabiensis*, 84% for *An. funestus* | [40] |
| Mosquito life expectancies | 18.64 days for *An. arabiensis* days, 21.5 days for *An. funestus* | Est. from [66] |
| Standard pyrethroid-only ITN resistance mortality in *An. funestus* | Homozygous: 10%, Heterozygous: 51.4% | [48] |
| Standard pyrethroid-only ITN resistance mortality in *An. arabiensis* | Homozygous: 23.3%, Heterozygous: 61.5% | [67,68] |
| pyrethroid-PBO ITN resistance mortality in *An. funestus* | Homozygous: 34%, Heterozygous: 67% | [69] |
| pyrethroid-PBO ITN resistance mortality in *An. arabiensis* | Homozygous: 36%, Heterozygous: 72% | Est. from [47,70] |
| IG2 ITN resistance mortality in *An. funestus* | Homozygous: 39%, Heterozygous: 78% | Est. from [71] |
| IG2 ITN resistance mortality in *An. arabiensis* | Homozygous: 41%, Heterozygous: 82% | Est. from [72] |
| IRS resistance mortality | Homozygous: 41.5%, Heterozygous: 83% | Est. from [47,68] |
| ITN and IRS useful lifetime | 3×365 days for ITN, 365 days for IRS | [14,20,58] |

## Results

We conducted a scenario analysis comprising two main simulation stages. First, we compared standard pyrethroid-only ITNs with the new generation ITNs (pyrethroid-PBO and IG2). Second, we evaluated the impact of changing from standard pyrethroid-only ITNs to the new bed nets on malaria transmission and the evolution of insecticide resistance. Each scenario was replicated across thirty independent stochastic realizations. Simulations were performed at resistance probabilities of 0.25, 0.5, and 0.75 (defined as the probability that an individual mosquito becomes resistant after exposure to pyrethroids). These values are illustrative rather than empirical and are intended to demonstrate the potential impact of interventions across different resistance levels.

### Impact of pyrethroid-PBO and IG2 ITNs on malaria transmission compared to standard pyrethroid-only ITNs

The simulation was conducted over a three-year period, with ITN deployment assumed to occur at the start of the peak malaria season in March (Figure 2). July, the peak transmission period, was used as the reference for evaluating ITN impact. The probability of mosquito resistance to pyrethroids was set at 0.5.

Figure 3 shows the impact of different ITN types on malaria incidence and prevalence in children under five. As shown in Figure 3(a), IG2 ITNs outperformed others, and also had sustained efficacy for up to 26 months, compared to 24 months for pyrethroid-PBO ITNs and 23 months for standard pyrethroid-only ITNs. By the end of year three, efficacy declined for all ITNs, but IG2 ITNs yielded the greatest reduction in incidence, followed by pyrethroid-PBO ITNs. In July (peak transmission) in the third year, standard pyrethroid-only ITNs resulted in 0.669 cases per person, while IG2 and PBO ITNs reduced incidence to 0.109 (83.7% reduction) and 0.265 (60% reduction), respectively. Figure 3(b) highlights similar trends in prevalence. By year three, although prevalence increased overall, IG2 ITNs showed the largest reduction. Prevalence with standard pyrethroid-only ITNs was 69.5%, compared to 34.7% with IG2 ITNs (50.1% reduction) and 47.7% with pyrethroid-PBO ITNs (31.3% reduction).

**Figure 3:**
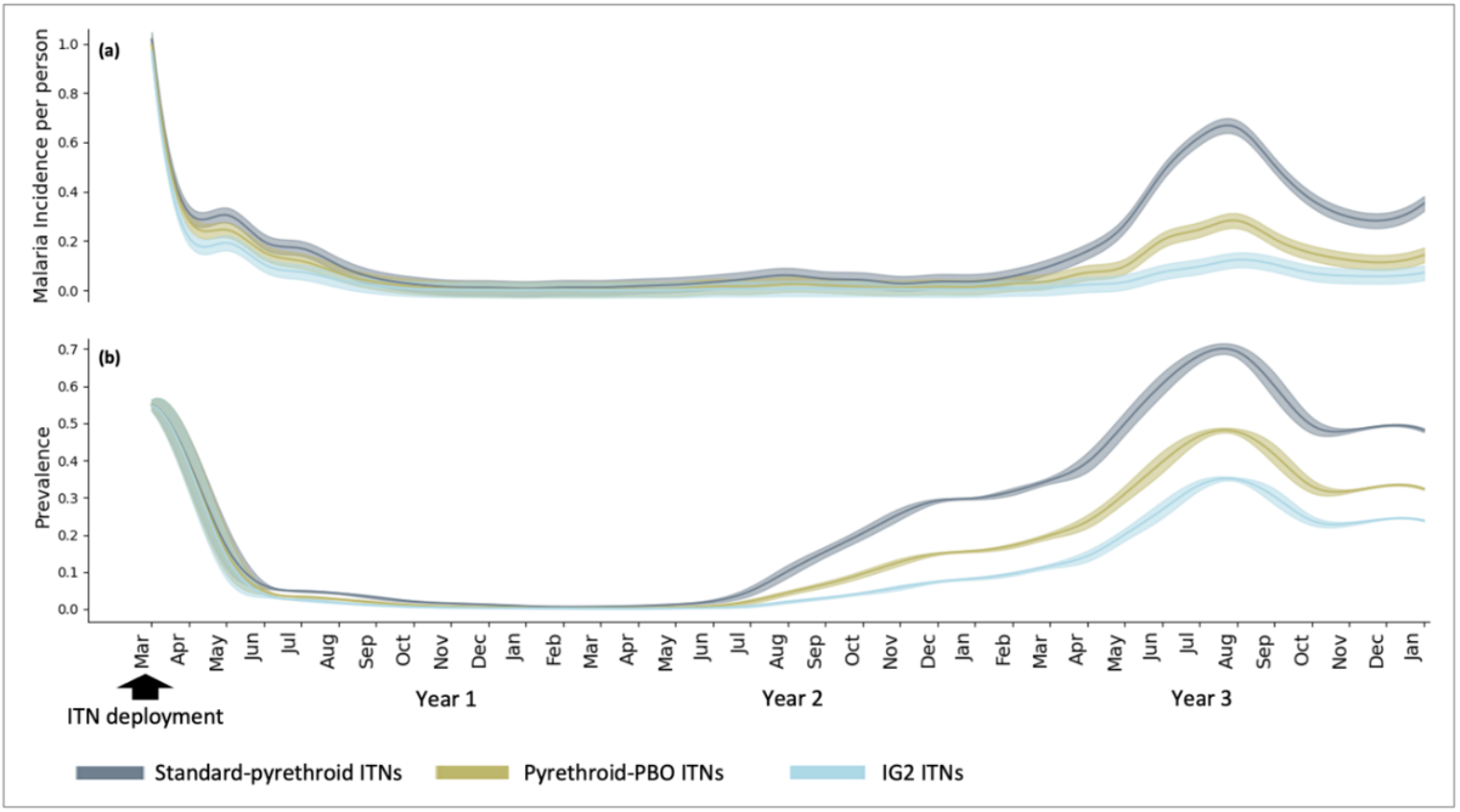
The impact of pyrethroid-PBO ITNs and IG2 ITNs on **(a)** malaria incidence and **(b)** prevalence in comparison with standard pyrethroid-only ITNs for children less than 5 years. Each type of ITN was assumed to be deployed at start of the peak malaria season in March.

Figure 4 presents the impact of ITNs on incidence and prevalence without age stratification. Unlike the findings for children under five, overall transmission intensity is lower. In July, the peak transmission period in the third year, IG2 ITNs reduced incidence and prevalence by 71% and 50.4%, respectively, compared to standard pyrethroid-only ITNs, while pyrethroid-PBO ITNs achieved reductions of 13.4% and 30.8%, respectively; emphasizing the superior long-term effectiveness of IG2 ITNs in reducing both malaria incidence and prevalence in high and seasonal-transmission settings.

**Figure 4:**
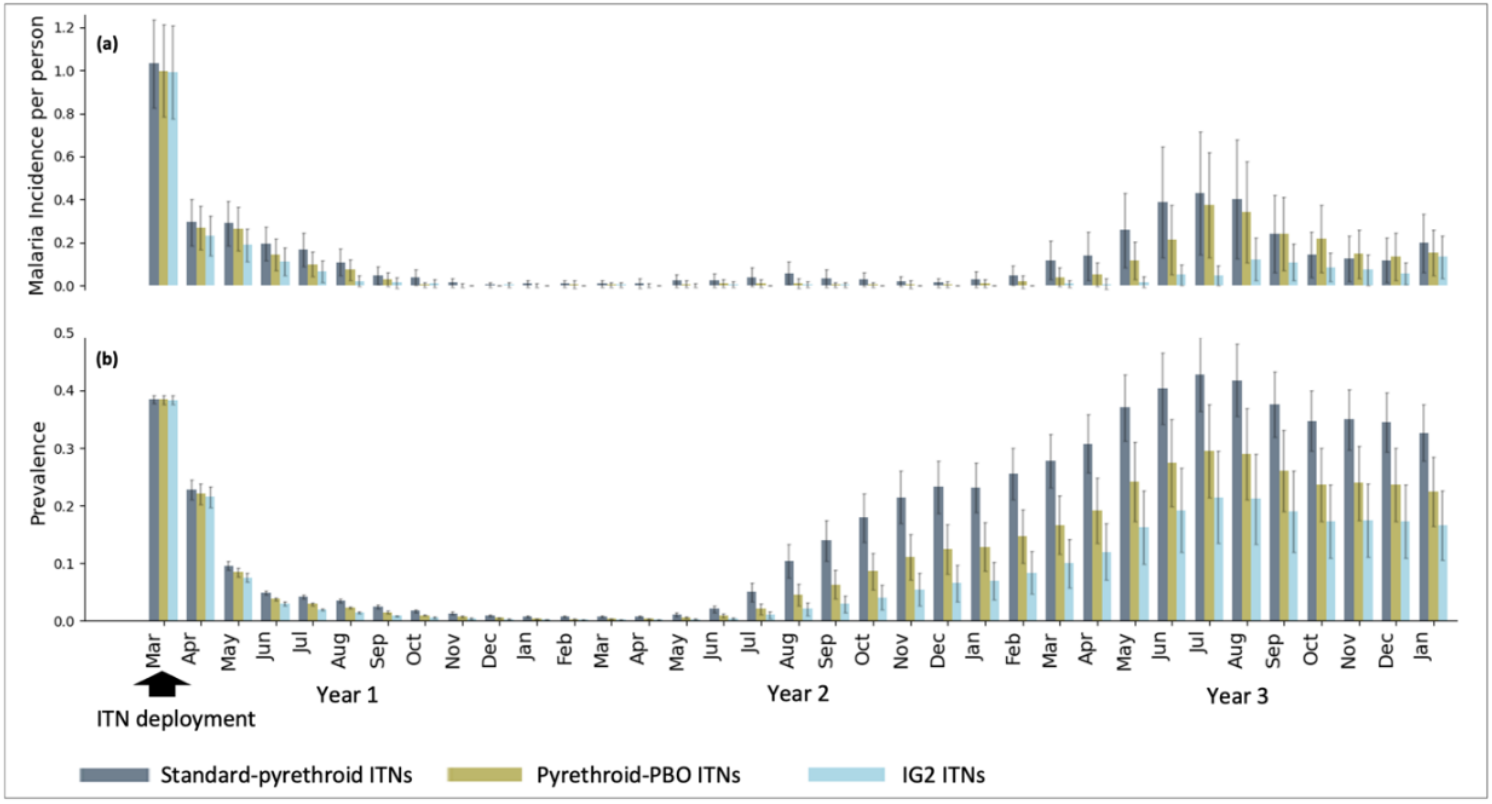
The impacts of pyrethroid-PBO ITNs and IG2 ITNs on **(a)** malaria incidence and **(b)** prevalence relative to standard pyrethroid-only ITNs without age-based stratification. All ITN types were assumed to be distributed at start of the peak malaria season in March.

Figure 5 and 6 illustrate the impact of the new ITNs on daily EIR relative to standard pyrethroid-only ITNs. As shown in Figure 6, IG2 ITNs reduced the EIR by 71% for *An. funestus* and 61% for *An. arabiensis* compared to standard pyrethroid-only ITNs in second year. In contrast, pyrethroid-PBO ITNs achieved reductions of 30% and 19% for *An. funestus* and *An. arabiensis*, respectively. Although IG2 ITNs outperformed other ITN types, EIR remained high in the third year across all ITN groups (Figure 5), highlighting the limitations of relying solely on ITNs for effective malaria control.

**Figure 5:**
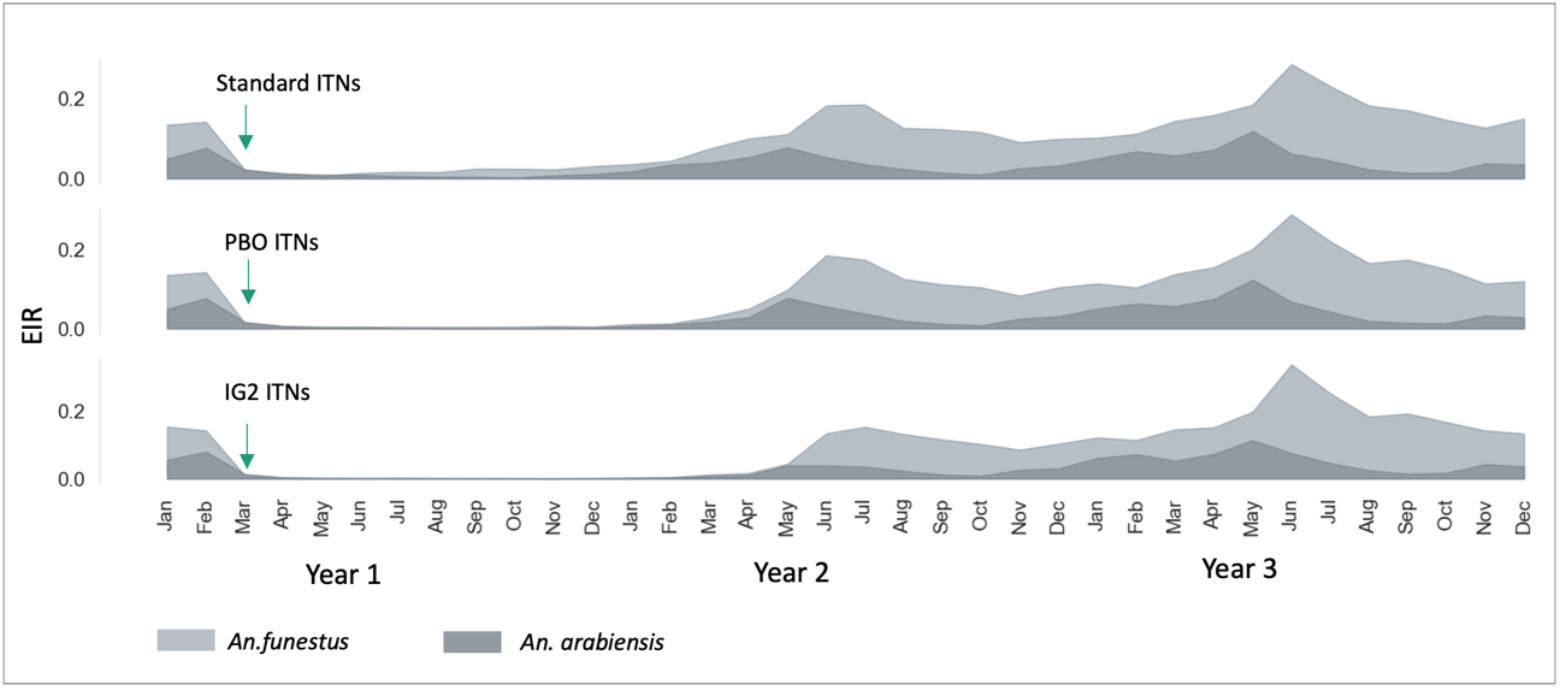
The impact of ITNs on daily EIR. *An. arabiensis* and *An. funestus* were assigned indoor feeding/resting fractions of 63.1% and 78.2% and anthropophilic indices of 62% and 84%, respectively [39,40]. Standard ITNs refer to pyrethroid-only ITNs, PBO ITNs to pyrethroid-PBO ITNs, and IG2 to Interceptor® G2.

**Figure 6:**
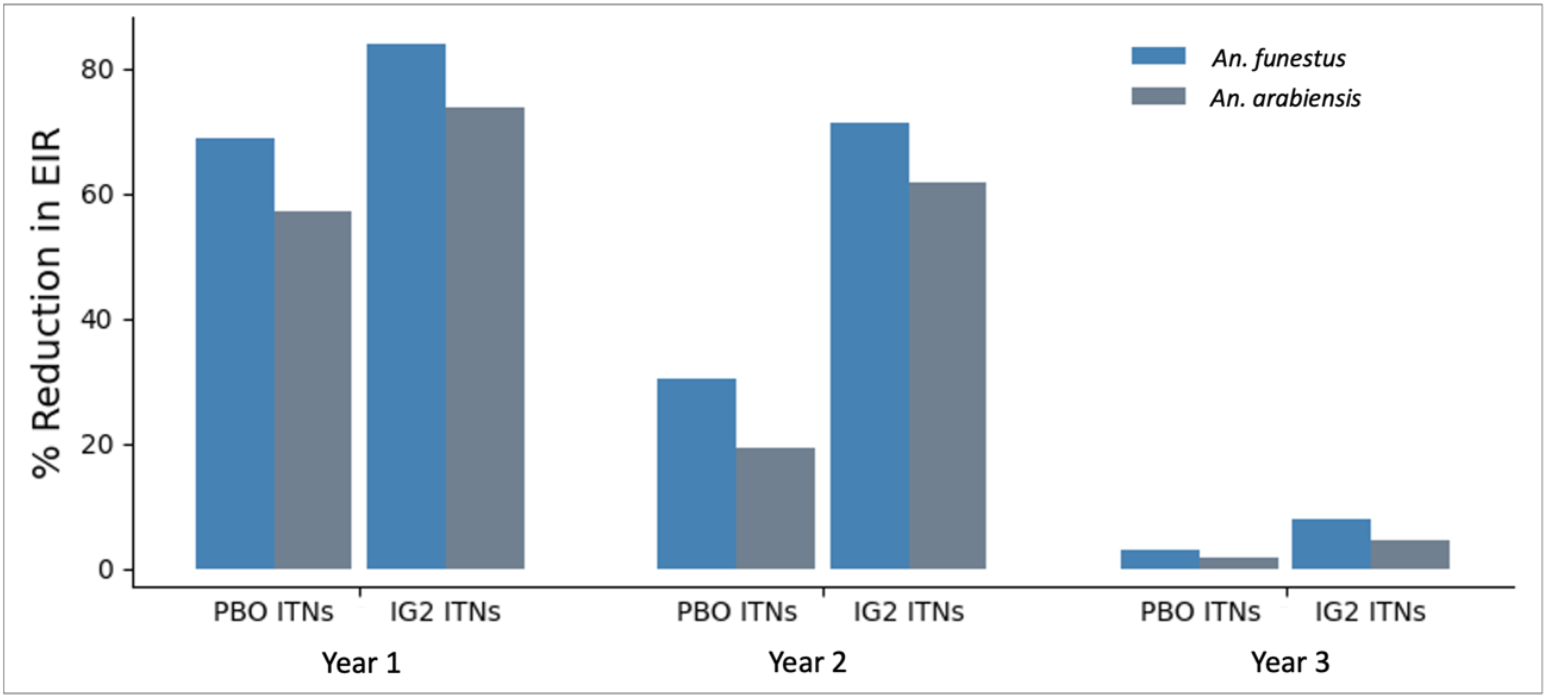
The percentage reduction in EIR relative to standard pyrethroid-only ITNs. PBO ITNs refer to pyrethroid-PBO ITNs, and IG2 to Interceptor® G2 ITNs.

### Impact of integrating IRS with pyrethroid-PBO ITNs

The impact of a single IRS implementation over a three-year period, implemented before the peak malaria transmission season in mid-May (at the end of rain season in the lake zone [45]), was simulated during the second or third year following the deployment of pyrethroid-PBO ITNs.

Figure 7 illustrates the impact of implementing IRS once every three years to mitigate the increase in malaria incidence and prevalence associated with the reduced efficacy of pyrethroid-PBO ITNs. When IRS is deployed before the peak malaria transmission season of the second year, the combined intervention reduces malaria incidence and prevalence by approximately 76.4% and 52%, respectively, in July of the third year. In contrast, when IRS is deployed before the peak malaria transmission season of the third year, malaria incidence and prevalence are reduced by approximately 72.4% and 51.4%, respectively, during the same period, compared to the strategy of deploying pyrethroid-PBO ITNs alone. Although both strategies show a resurgence of cases in the third year, deploying IRS in the second year offers a more sustained reduction in malaria burden and outperforms the strategy of deploying IRS in the third year.

**Figure 7:**
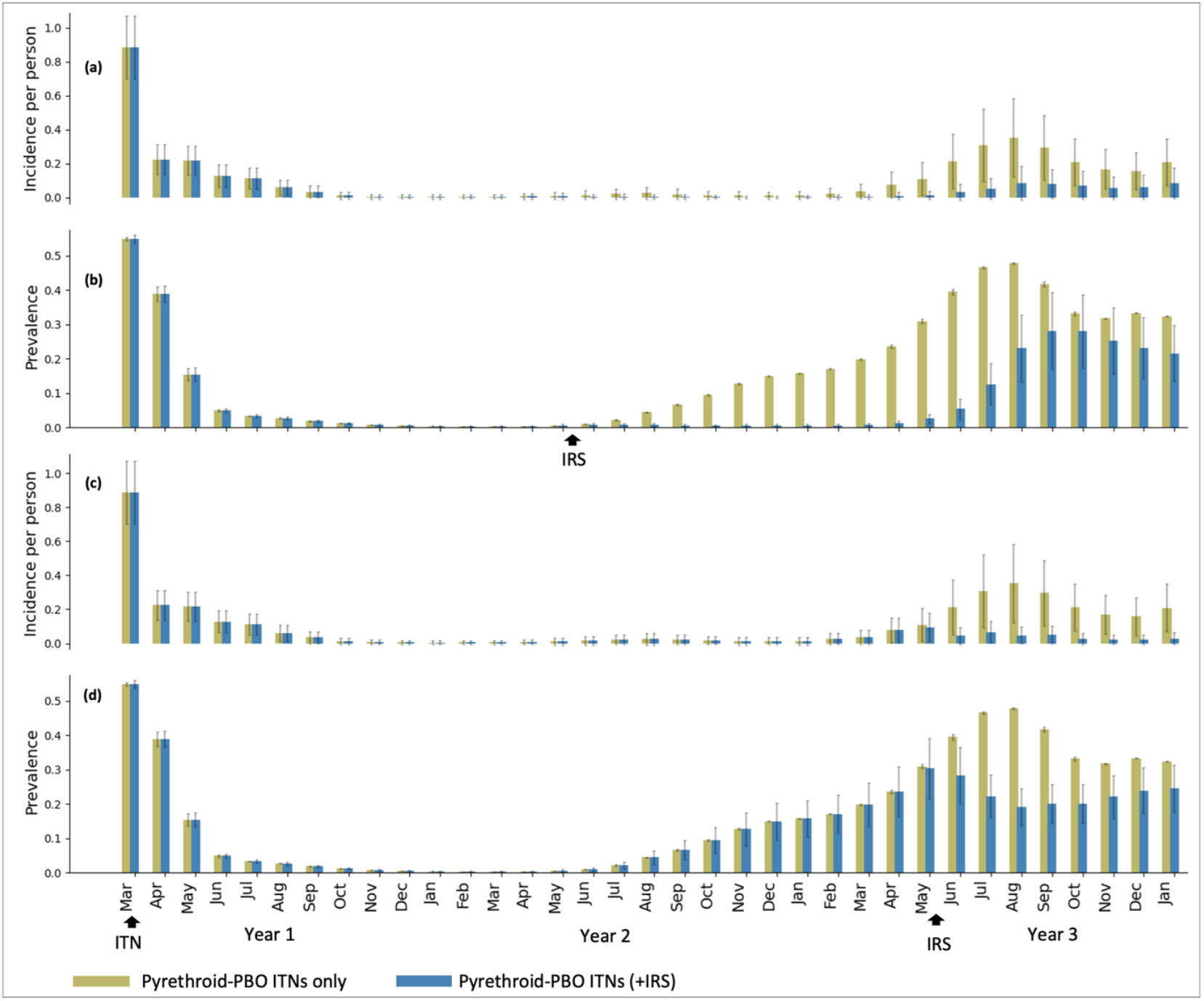
Impact of a single IRS round every three years on malaria incidence and prevalence among children under five. Panels (a) and (b) show IRS deployed before the peak transmission season in mid-May of the second year, while panels (c) and (d) show deployment in the third year. ITNs were assumed to be distributed at the start of the peak transmission season in March, with IRS scheduled in mid-May after the rainy season [45].

### Impact of changing from standard pyrethroid-only nets to new types of ITNs

To illustrate the selection dynamics of resistant mosquitoes under ITNs transition versus continuous deployment of standard pyrethroid-only ITNs, resistance is modeled through genotype classifications: homozygous susceptible (*a*_0_: *a*_0_), heterozygous (*a*_0_: *a*_1_), and homozygous resistant (*a*_1_: *a*_1_). Heterozygous mosquitoes are assumed to be partially resistant, while homozygous resistant and susceptible mosquitoes are considered fully resistant and susceptible, respectively [25,46–48]. Simulations were run under three phenotypic resistance probabilities (0.25, 0.5 and 0.75), to assess effects on vector selection, and in turn, malaria incidence and prevalence. Additionally, four ITN deployment strategies were simulated over nine years, each deployment comprising three-year blocks with varying ITN types.

#### Repeated deployments of standard pyrethroid-only ITNs

Figure 8(a) shows the impact of repeated deployment of standard pyrethroid-only ITNs over nine consecutive years. Continued application of ITNs with the same insecticidal mode of action exerts strong selection pressure on mosquito populations, promoting resistance. At a resistance probability of 0.75, homozygous susceptible (*a*_0_: *a*_0_) mosquitoes decline sharply in the first three years due to pyrethroid-induced mortality, while homozygous resistant (*a*_1_: *a*_1_) mosquitoes increase gradually while the heterozygous (*a*_0_: *a*_1_) mosquitoes display intermediate survival. In the next three years, ongoing ITN use intensifies selection, further increasing *a*_1_: *a*_1_ frequencies, with a decline in *a*_0_: *a*_0_ and *a*_0_: *a*_1_. By year nine, *a*_1_: *a*_1_ frequencies reach 0.704 in *An. funestus* and 0.502 in *An. arabiensis*, reflecting strong, sustained selection. This pressure weakens as resistance probability decreases. Additionally, the prolonged deployment of the same ITNs sustains malaria burden, as shown in Figure 9 (red curve), with incidence changing from 0.48 in July of the first year to 0.47 in the third year, and prevalence from 0.70 to 0.677.

**Figure 8:**
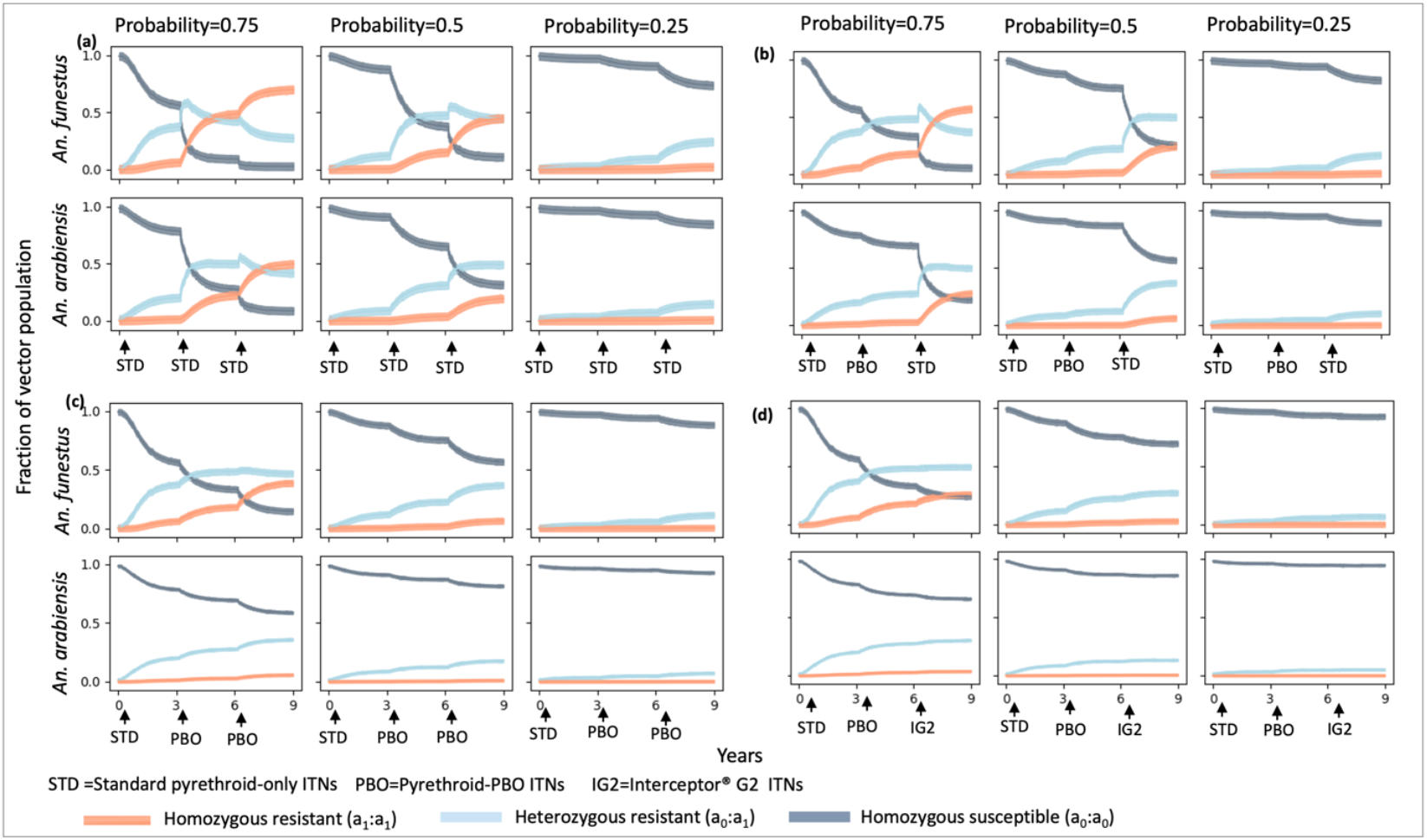
The impact of changing ITNs on vector genetics compared to repeated use of standard pyrethroid-only ITNs. Columns (probabilities 0.75, 0.5, and 0.25) represent scenarios corresponding to the probability of mosquitoes surviving pyrethroid exposure. Additionally, *a*_0_: *a*_0_ and *a*_1_: *a*_1_ denote the proportions of mosquitoes with homozygous sensitive and resistant allele combinations, respectively, while *a*_0_: *a*_1_ represents the proportion of mosquitoes with heterozygous resistant allele combinations.

**Figure 9:**
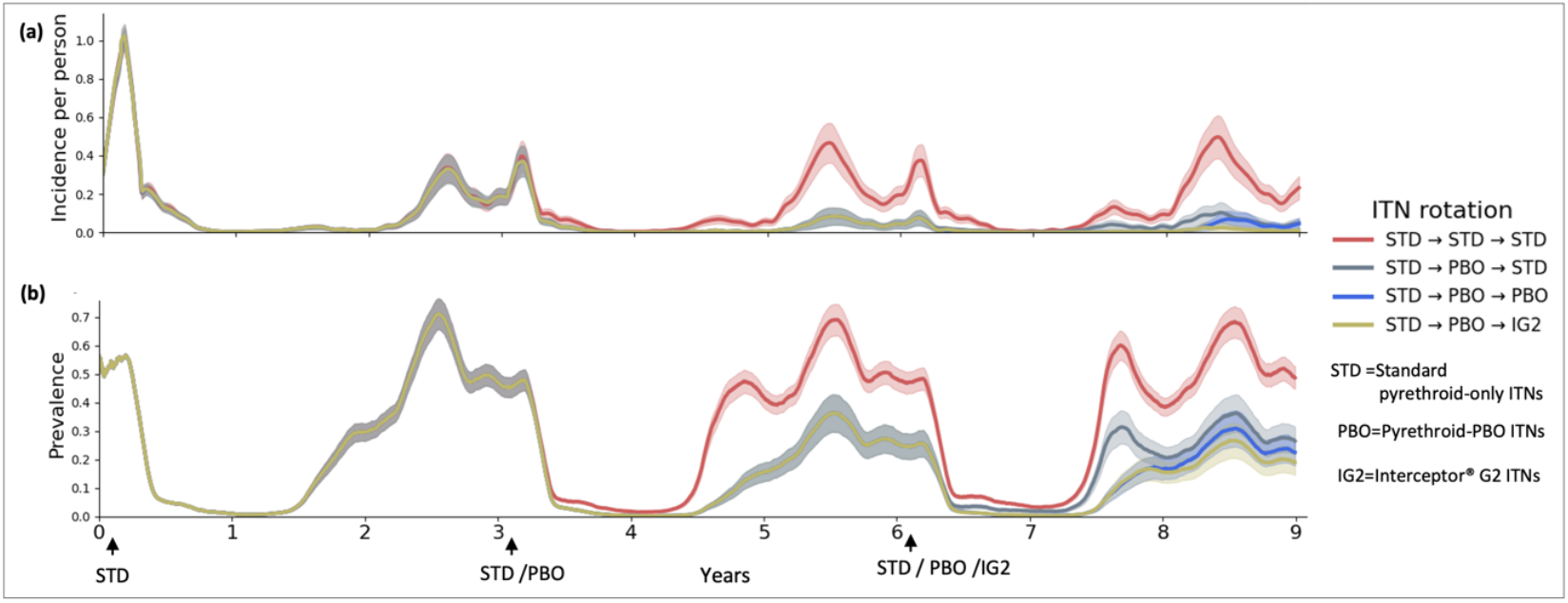
The impact of ITN transition: **(a)** incidence **(b)** prevalence for children under five, compared to repeated deployments of standard pyrethroid-only ITNs. ITNs are distributed triennially, commencing at the at start of the peak malaria season on March, each at 80% coverage when probability of a mosquito being resistant to pyrethroids killing is set to be 0.5.

#### Rotational deployments of standard pyrethroid-only ITNs and pyrethroid-PBO ITNs

Figure 8(b) illustrates the impacts of rotating between standard pyrethroid-only and pyrethroid-PBO ITNs. During the initial phase, similar selection trends to those in Figure 8(a) are observed. The switch to pyrethroid-PBO ITNs reduces this pressure, leading to a continued decline in homozygous susceptible (*a*_0_: *a*_0_), a marginal increase in homozygous resistant (*a*_1_: *a*_1_) and heterozygous (*a*_0_: *a*_1_) mosquitoes. Reintroduction of standard pyrethroid-only ITNs in the final three years results in a further drop in *a*_0_: *a*_0_, a substantial rise in *a*_1_: *a*_1_, and a modest decline in *a*_0_: *a*_1_. By year nine, this rotation strategy reduces *a*_1_: *a*_1_ frequencies by 18.5% (*An. funestus*) and 83% (*An. arabiensis*) at resistance probability 0.75, and by 44.5% and 69.3%, respectively, at probability 0.5, compared to continuous standard pyrethroid-only ITN use (Figure 8(a)). As shown in Figure 9 (grey graph), this approach also lowers malaria incidence and prevalence by 77% and 48.6% respectively.

#### Transitioning from standard pyrethroid-only ITNs to pyrethroid-PBO ITNs

Figure 8(c) depicts the impact of deploying the standard pyrethroid-only ITNs for three years followed by pyrethroid-PBO ITNs for six years. Selection patterns during the initial standard pyrethroid-only ITN phase mirror those observed in Figure 8(a) and (b). Changing to pyrethroid-PBO ITNs reduces selection pressure on homozygous resistant (*a*_1_: *a*_1_) mosquitoes, even at higher resistance probabilities. By year nine, this strategy lowers *a*_1_: *a*_1_ frequencies by 46.4% (*An. funestus*) and 85.5% (*An. arabiensis*) at resistance probability 0.75, and by 82.1% and 91.7%, respectively, at probability 0.5, compared to continuous standard pyrethroid-only ITN use. However, prolonged use of pyrethroid-PBO ITNs may still drive selection pressure for resistance over time, particularly in regions where *An. funestus* predominates. As shown in Figure 9 (blue graph), this transition also reduces malaria incidence and prevalence by 87% and 62% respectively.

#### Transitioning from standard pyrethroid-only ITNs to pyrethroid-PBO ITNs and then IG2 ITNs

Figure 8(d) shows the impact of transition from standard pyrethroid-only ITNs to pyrethroid-PBO ITNs and then IG2 ITNs. Similar selection trends to those in Figure 8(a), (b) and (c) are observed during the initial standard pyrethroid-only ITN deployment phase. The subsequent deployment of pyrethroid-PBO and IG2 ITNs, reduces selection pressure and limits resistance development. Compared to repeated standard pyrethroid-only ITN deployments (Figure 8(a)), this strategy reduces homozygous resistant (*a*_1_: *a*_1_) frequencies by 62.2% (*An. funestus*) and 92.8% (*An. arabiensis*) at resistance probability 0.75, and by 93.1% and 94.1%, respectively, at probability 0.5. Additionally, as shown in Figure 9 (olive-like curve), this strategy also lowers malaria incidence and prevalence by 94% and 75.2% respectively.

Across all scenarios, homozygous resistant *An. funestus* showed consistently stronger selection than *An. arabiensis*. The strategy of transitioning from pyrethroid-only ITNs to pyrethroid-PBO and then IG2 ITNs is most effective in reducing selection pressure, incidence and prevalence. The results presented here provide significant public health implications, offering valuable guidance for managing insecticide resistance and malaria transmission more effectively. Specifically, they emphasize that understanding the resistance profile of mosquito populations is essential for assessing the impact of ITNs on resistance dynamics and for informing strategies to minimize resistance development while maintaining the efficacy of vector control interventions.

## Discussion

Malaria transmission in many areas in Tanzania is seasonal and characterized by high levels of insecticide resistance [35]. However previous models investigating the impacts of these trends on malaria control have often overlooked the interactions of key factors such as multiplicity of important vector species, associated species-specific vector dynamics, seasonality and resistance selection, instead investigating them in isolation [24–28]. In this study, we employed the EMOD modeling platform to capture seasonal transmission patterns, insecticide resistance, and the population dynamics of *An. funestus* and *An. arabiensis*. We evaluated the impact of pyrethroid-PBO and IG2 ITNs, in comparison with standard pyrethroid-only ITNs and their sequential deployment strategies, on malaria incidence, prevalence, and the spread of insecticide resistance.

In settings where malaria transmission is both seasonal and endemic [35,49], deploying interventions at the onset of the peak transmission season is considered the most effective strategy for maximizing their impact [26,50]. The agent-based model used in this study demonstrates that deploying ITNs at start of the peak malaria season in March, optimizes their effectiveness. It also highlights the superior effectiveness of IG2 ITNs in reducing malaria transmission compared to pyrethroid-PBO and standard pyrethroid-only ITNs. In alignment with the deployment of pyrethroid-PBO nets across all epidemiological strata in Tanzania [19,23], the current study reveals that pyrethroid-PBO ITNs demonstrate significantly greater effectiveness than pyrethroid-only ITNs, although their efficacy is lower compared to IG2 ITNs. The enhanced performance of pyrethroid-PBO ITNs can be partly attributed to the synergistic action of pyrethroid-PBO in mitigating pyrethroid resistance, aligning with findings from other epidemiological studies conducted in various locations [51,52].

The superior efficacy of dual active-ingredient ITNs compared with standard pyrethroid-only ITNs against major malaria vectors has been well documented [37,53–55], with most studies reporting a greater impact on *An. funestus* than on other vector species. In line with these findings, the present study demonstrates that IG2 ITNs can substantially reduce the EIR of *An. funestus* relative to *An. arabiensis*. However, despite their improved performance over other ITN types, EIRs remained high across all intervention arms by the third year, highlighting the limitations of ITNs alone and the necessity of integrated vector management for sustained malaria control and elimination. The findings also underscore the influence of new-generation ITNs on insecticide resistance dynamics in *An. funestus* and *An. arabiensis*. Consistent with prior evidence identifying *An. funestus* as a dominant vector in high-resistance settings [34,35,56], our modeling indicates stronger selection for *An. funestus* than *An. arabiensis*, reinforcing the need for strategic deployment of other interventions such as novel ITN formulations in such settings to mitigate resistance and reduce transmission.

Although long-lasting IRS insecticides are available [57], implementation is hindered by high costs and the disruptive nature of annual applications [58]. Consequently, IRS has been phased out in Tanzania in favor of pyrethroid-PBO ITNs [22] despite limited evidence supporting their ability to fully replace IRS in high-transmission and resistance areas [22]. While transitioning from standard pyrethroid-only ITNs to pyrethroid-PBO ITNs can avert twice as many cases and help manage resistance [26], their efficacy wanes significantly within two years [51]. This study shows that combining pyrethroid-PBO ITNs with IRS applied once every three years can effectively curb malaria resurgence due to declining ITN efficacy, sustaining low transmission. Specifically, the model indicates that implementing IRS in the second year of each three-year cycle provides a more durable reduction in malaria burden, representing an optimal strategy for the NMCP to accelerate progress toward the goal of a malaria-free Tanzania [19]. Despite concerns about the feasibility of combined use in some settings [14,59], evidence supports substantial health benefits from integrating IRS with ITNs [59,60]. The shifting IRS from annual to triennial application may offer a cost-saving advantage. Although triennial IRS strategy has not yet been implemented in practice, field studies are recommended to assess its feasibility and effectiveness.

The repeated use of insecticides with the same mode of action contributes substantially to the development of resistance in mosquito populations [61,62]. Consistent with previous findings [3,13,25,34], the current study shows that continuous deployment of standard pyrethroid-only ITNs imposes strong selection pressure, promoting resistance alleles and reducing ITN efficacy over time. This stresses the limitations of relying solely on standard pyrethroid-only ITNs for sustained malaria control. The presented model emphasizes the critical importance of introducing new types of ITNs in mitigating resistance and lowering malaria incidence and prevalence. Transition and rotating ITNs with different insecticidal modes of action, such as pyrethroid-only, pyrethroid-PBO and IG2 ITNs, interrupts resistance selection, decreases resistant allele frequencies, and enhances transmission control. Notably, including IG2 ITNs in transition yields the greatest reduction in both resistance development and malaria burden. Implementing this strategy in high-resistance and transmission areas like the lake zone Tanzania is essential for achieving long-term malaria reduction and advancing elimination efforts.

This study contributes to the growing body of evidence supporting malaria elimination efforts in Tanzania. However, the model simulates malaria transmission only for *Plasmodium falciparum*, excluding other species; this limitation is unlikely to materially affect the results, as *Plasmodium falciparum* accounts for over 96% of malaria infections in mainland Tanzania [19]. Additionally, the study focuses exclusively on ITN-related insecticide resistance, excluding other sources such as agricultural insecticide use; thus, findings should be interpreted within the study’s assumptions and with caution when generalizing without regional calibration using local entomological and epidemiological data. To our knowledge, this is the first study to assess novel ITNs in the context of insecticide resistance, incorporating seasonality and quantifying dynamics of both *An. funestus* and *An. arabiensis*, while also predicting resistance development to new insecticide classes, an aspect previously unaddressed.

## Conclusion

The effective malaria control in Tanzania, particularly in high-transmission zones and resistant areas, requires integrated, and adaptive vector control tools and strategies. The study indicates that continuous use of pyrethroid-only ITNs maintains strong selection pressure and reduces the impact of these interventions on malaria transmission. In contrast, transitioning or rotating ITNs with diverse modes of action, together with periodic use of IRS, effectively mitigates insecticide resistance and supports long-term malaria control.

## Data Availability

All data supporting this study are included within the manuscript

## Abbreviations

ITNs: Insecticide-treated nets
EMOD: Epidemiological modeling software
IRS: Indoor residual spraying
WHO: World Health Organization
NMCP: National Malaria Control Programme

## Appendix 1

### Description on how the homozygous and heterozygous genotype values were estimated

The estimated mortalities corresponding to heterozygous genotype frequencies for ITNs and IRS were calculated using Equation (1), adapted from WHO guidelines [10], with homozygous mortality assumed to be half that of heterozygous mortality. The estimated values represent the genotype-specific mortality for the interventions and were used as killing modifiers in EMOD to model resistance in homozygous and heterozygous mosquitoes. Although derived from experimental or field data, these estimates are context-specific approximations, should not be considered universal constants or definitive guidelines.

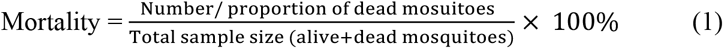

## Acknowledgements

The authors also acknowledge the Institute for Disease Modelling (IDM) for providing access to the EMOD platform. The authors sincerely thank the National Malaria Control Programme (NMCP) of Tanzania for granting access to the incidence data used for model calibration. Gratitude is also extended to the Outdoor Mosquito Control (OMC) group at the Ifakara Health Institute (IHI) for their valuable contributions to the development of this manuscript.

## Author Contributions

H.E.G., Y.P.M., H.S.N., F.O.O., and P.S. conceived and designed the study. H.E.G. conducted all simulations with guidance from P.S., and drafted the initial manuscript and subsequent revisions. M.M.M., F.C., Y.P.M., H.S.N., F.O.O., and P.S. supervised the study and critically reviewed and edited the manuscript drafts. All authors revised the final manuscript, and approved its submission.

## Availability of Data and Materials

All data supporting this study are included within the article.

## Declarations

### Ethical approval and consent to participate and publish

Ethical approval for this study was obtained from the Institutional Review Board (IRB) of the Ifakara Health Institute (IHI/IRB/EXT/No: 44-2024) and from the Medical Research Coordination Committee of the National Institute for Medical Research (NIMR), Tanzania (NIMR/HQ/R.8c/Vol.I/2937). Consent to publish was obtained from NIMR (Ref. No. BD.242/437/01C/184).

